# Age-standardised trends in Incidence Rates of Noncommunicable diseases among Adults Aged 30 to 79 in Senegal from 2000 to 2019

**DOI:** 10.1101/2025.09.26.25336768

**Authors:** Ngone Diaba Gaye, Mohamed B. Jalloh, Tiffany L. Gary-Webb, Mame Madjiguene Ka, Malick Anne, Ishta Madan, Ahmed Tamer Abdelnoor, John Tukakira, Damaris Kyem, Gurbinder Singh, Jennifer L. Carter, Elisabeth L.P. Sattler, Modou Jobe, Bamba Gaye

## Abstract

**Introduction:** Non-communicable diseases (NCDs) represent a significant global health burden in sub-Saharan Africa (SSA). However, most SSA countries such as Senegal lack reliable data despite the need for surveillance of NCD trends to inform targeted preventive strategies.

**Methods:** We used publicly available data from the Global Burden of Disease Study (World Health Organization’s Health Inequality Data Repository). We used age-standardised regression to analyze the trends of NCDs (cancers, cardiovascular diseases, chronic respiratory diseases, diabetes, and chronic kidney diseases) incidence among adults in Senegal. Statistical analysis was performed using the Joinpoint Regression Program, Version 5.3.0.0 and data visualization was performed using R, version 4.4.0

**Results:** NCD incidence rates in females were consistently higher than in males. Rates in females increased from 190,857 to 194,620 cases per 100,000 population between 2000 and 2019 (p-value 0.007) as compared to 160,623 to 163,800 cases per 100,000 population in males (p-value 0.002). Incidence rates were higher in females for neoplasm (p-value 0.03), chronic respiratory diseases (p-value 0.07), and diabetes and chronic kidney diseases (p-value 0.04), while cardiovascular disease incidence was higher in males (p-value 0.02).

**Conclusion:** There was a significant increase in age-standardized NCD incidence among adults in Senegal. When considering subtypes, all had increased incidence rates, except cardiovascular diseases which showed a significant decrease over time. A national NCD registry and targeted risk factor prevention programs are crucial to help tackle the NCD burden in Senegal.

**KEY MESSAGES:** *What is already known?:*

- Senegal is experiencing a shift towards a higher burden of NCD, with a probability of dying from NCD as high as 20% [95% CI: 11.9-28.8%] in 2019 according to WHO.
- Unfortunately, reliable epidemiological data and surveillance systems are lacking to inform appropriate prevention and resource allocation strategies.

*What does this study add?:*

- Overall, we observed a significant increasing trend in the age-standardized incidence of NCD during the study period, 2000-2019.
- There were differences between NCD subtypes, with neoplasm rates remaining constant throughout the study period while diabetes, chronic kidney disease and chronic respiratory disease were increasing.
- Incidence rates were consistently higher in females compared to males, especially for chronic respiratory disease, diabetes, and chronic kidney disease.

*What do the new findings imply?:*

- The overall non-significant increasing trend of NCD incidence rates in Senegal calls for the critical need to invest in robust surveillance systems such as NCD national registry and targeted risk factors prevention programs to tackle this growing burden.
- Discrepancies in the trends of some NCD subtypes may reflect reporting bias, which needs to be addressed by well-designed longitudinal cohort studies.

## INTRODUCTION

Non-communicable diseases (NCDs), including cardiovascular diseases, cancer, chronic respiratory diseases, and diabetes, account for most deaths worldwide, and their burden is rising fastest in low- and middle-income countries (LMICs).^1^ Reliable data from LMICs are essential for prevention, service planning, and policy, yet many countries in sub-Saharan Africa (SSA) have limited vital registration, weak routine surveillance, and NCD indicators that are poorly integrated into national health information systems.^1^

Senegal illustrates these challenges. Limited evidence from national risk factor surveys and facility-based series suggests increasing prevalence hypertension, diabetes, and selected cancers,^2–8^ alongside exposure to tobacco use, physical inactivity^9^ and diets high in sugars, salt, and saturated fats.^10^ These changes plausibly reflect population growth and rapid urbanization.^6^ According to the World Health Organization (WHO), NCDs were estimated to be the leading broad cause of death in Senegal in 2019 and the probability of dying from NCD was as high as 20% [95% CI: 11.9-28.8%].^11^

Structural barriers compound this mortality context, including late presentation, ^7,12^ rural–urban disparities in access, limited financial protection, constrained medicine availability, and suboptimal adherence.^7,13,14^ Weak integration of NCD data into the national health information system remains a critical challenge, leading to fragmented and incomplete surveillance. Weak mortality and morbidity reporting, limited use of electronic health records, and inadequate funding for data infrastructure hinder effective disease monitoring and policy development. To tackle the burden of NCD, governments need surveillance systems and infrastructure to produce reliable data that can direct policies and legislations that will ultimately prevent the burden of NCD.^1,15^

Timely, population-based estimates of incidence are needed to guide prevention and service delivery. We therefore examined age-standardised incidence rates (ASIR) among adults in Senegal from 2000 to 2019. We hypothesized that ASIR increased over time, reflecting demographic change, urbanization, and rising exposure to modifiable risk factors, with the aim of informing national strategies for NCD prevention, early detection, and treatment.

## MATERIALS AND METHODS

### Study design and data sources

We conducted a secondary analysis of ASIR for Senegal using estimates from the Global Burden of Disease (GBD) 2019 study that we accessed via the WHO Health Inequality Data Repository (HIDR).^16^ HIDR is a public catalogue of disaggregated health datasets (launched April 2023); some series originate from external sources and are not “WHO official estimates.” In our case, all ASIRs were Institute for Health Metrics and Evaluation (IHME)/GBD-2019 outputs retrieved through HIDR.

### Case definitions and cause hierarchy

Outcomes followed the GBD cause hierarchy: Level-1 NCDs and Level-2 categories neoplasms, cardiovascular diseases, chronic respiratory diseases, and diabetes and kidney disease.

GBD obtained incidence estimates by using a Bayesian meta-regression to synthesise several input sources, including surveys, censuses, vital statistics, and other health-related data sources.^17^ For non-fatal outcomes, DisMod-MR 2.1 (GBD Bayesian disease model meta-regression platform) produces internally consistent incidence, prevalence, remission, and excess mortality estimates; cause-specific deaths are estimated with CODEm (Cause of Death Ensemble model). NCDs refer to noninfectious diseases^17^. Neoplasms refer to abnormal tissue growths, both benign and malignant.^17^ Cardiovascular diseases are a group of disorders of the heart and blood vessels and include coronary heart disease, cerebrovascular disease, rheumatic heart disease, and other conditions^17^. Chronic respiratory diseases affect the airways and other structures of the lungs and include chronic obstructive pulmonary disease, asthma, occupational lung diseases and pulmonary hypertension.^17^

### Senegal input data and provenance

To enhance transparency, we enumerated all Senegal-specific input series contributing to these estimates (e.g., population-based surveys, registries, health-service data) using the GBD 2019 Data Input Sources tool. Where local data are sparse, GBD models borrow strength across space and time via a hierarchical “geographical cascade” and spatiotemporal Gaussian process regression, which we note as a limitation for country-level precision.

ASIR were directly standardised to the GBD world standard population using 5-year age groups (both sexes, all ages) and reported per 100,000 population with 95% confidence intervals (CIs). This differs from WHO 2000–2025 weights; readers should interpret cross-study comparisons accordingly. We analysed the four quinquennial time points available in HIDR for Senegal (2005, 2010, 2015, 2019).

### Statistical analysis

We used the Joinpoint Regression Program, Version 5.3.0.0 to analyze data and R, version 4.4.0 (R Foundation for Statistical Computing) for data visualization. We analyzed statistical significance of trends using an annual percent change (APC) and report the results.^18^ Joinpoint regression uses the grid search method and segmented regression to model data.^19^ It is able to handle correlated data by allowing specification of variance-covariance matrices to specify correlated data. Standard errors used in the analysis were back calculated from confidence intervals in the datasets using the method described by Julian PT Higgins and others.^20^

### Ethics, data, and code availability

All data are publicly available (HIDR and IHME/ Global Health Data Exchange [GHDx]). No individual-level identifiers are present; institutional ethics review was not required. Analysis code and tidy datasets will be deposited in an open repository upon acceptance, and URLs to the HIDR dataset(s) and GBD query will be provided.

## RESULTS

Overall, we found an increasing trend in NCD ASIR. Subgroup analysis revealed variations in ASIR trends by sex. Incidence rates in females were consistently higher than in males. Rates in females increased from 190,857 cases per 100,000 population [95% CI 180,864-201,390] in 2000 to 194,620 cases per 100,000 population in 2019 [95% CI 184,482-204,560] (p-value 0.007) with a 0.17 annual percent increase. In males, NCD ASIR increased from 160,623 cases per 100,000 population [95% CI 151,716-170,325] in 2000 to 163,800 cases per 100,000 population in 2019 [95% CI 155058-173,439] (p-value 0.002) with a 0.11 annual percent increase.

There were differences in the trends by type of NCD. Rates of neoplasms were consistently higher in females compared to males (**Figure 1**). The rate of neoplasms increased in females from 3,323 cases per 100,000 population [95% CI 2,705-4,093] in 2000 to 3,351 cases per 100,000 population [95% CI 2,722-4,130] in 2019 (p-value 0.03) with a 0.04 annual percent increase while it also increased slightly in males from 3,118 cases per 100,000 population [95% CI 2,562-3,806] in 2000 to 3,148 cases per 100,000 population [95% CI 2,593-3,838] in 2019 (p-value 0.03) with a 0.04 annual percent increase.

**Figure 1:**
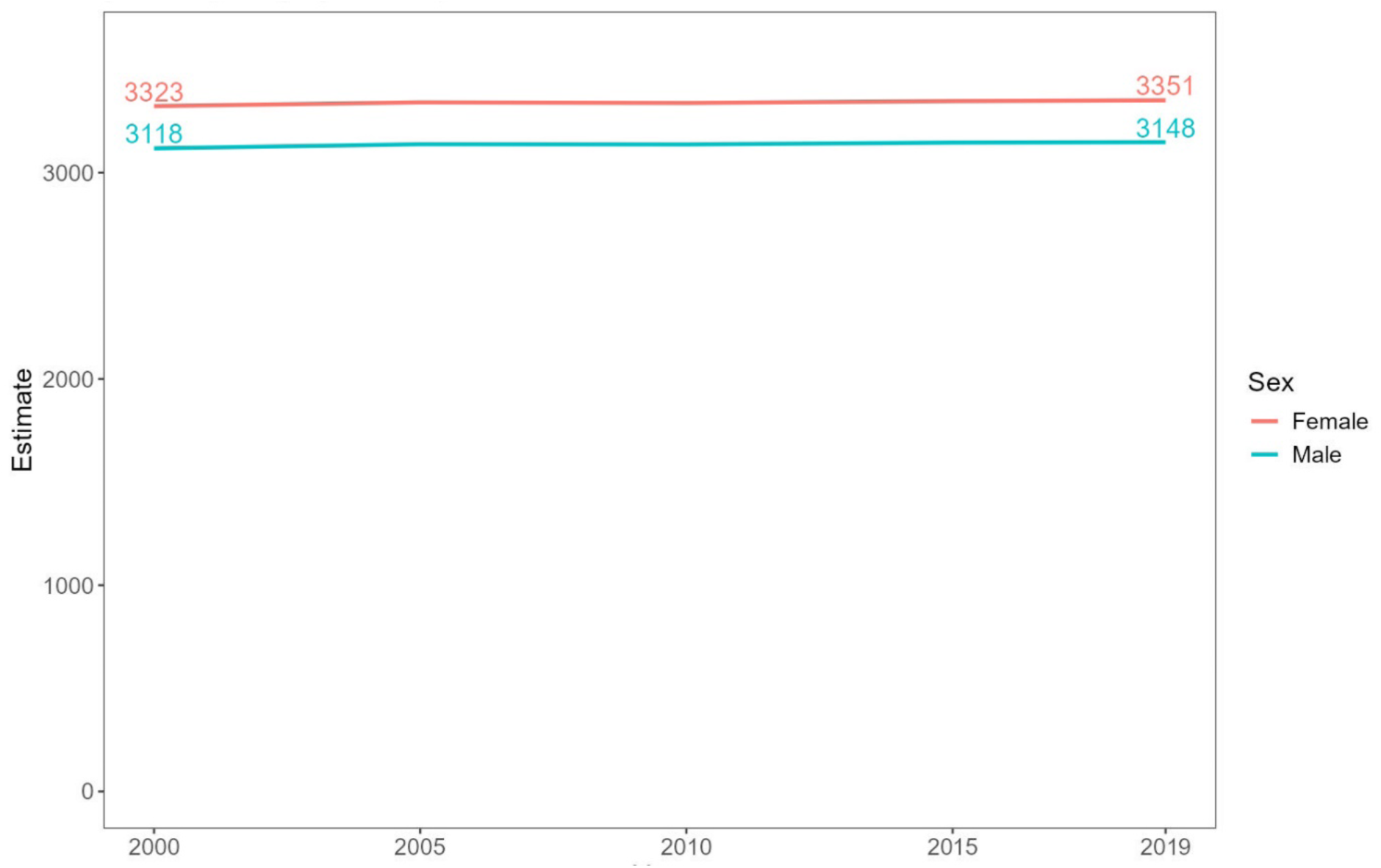
Neoplasms ASIR by sex (per 100 000 population), stable over time (p-value for trend 0.08) and consistently higher in women.

(**Figure 2**). Rates of cardiovascular disease were consistently higher in males compared to females. Incidence in males decreased from 652 cases per 100,000 population [95% CI 608-701] in 2000 to 631 cases per 100,000 population [95% CI 587-677] in 2019 (p-value 0.02) with a 0.18 annual percent decrease while it decreased from 598 cases per 100,000 population [95% CI 561-640] in 2000 to 584 cases per 100,000 population [95% CI 546-623] in 2019 in females (p-value 0.02) with a 0.12 annual percent decrease.

**Figure 2:**
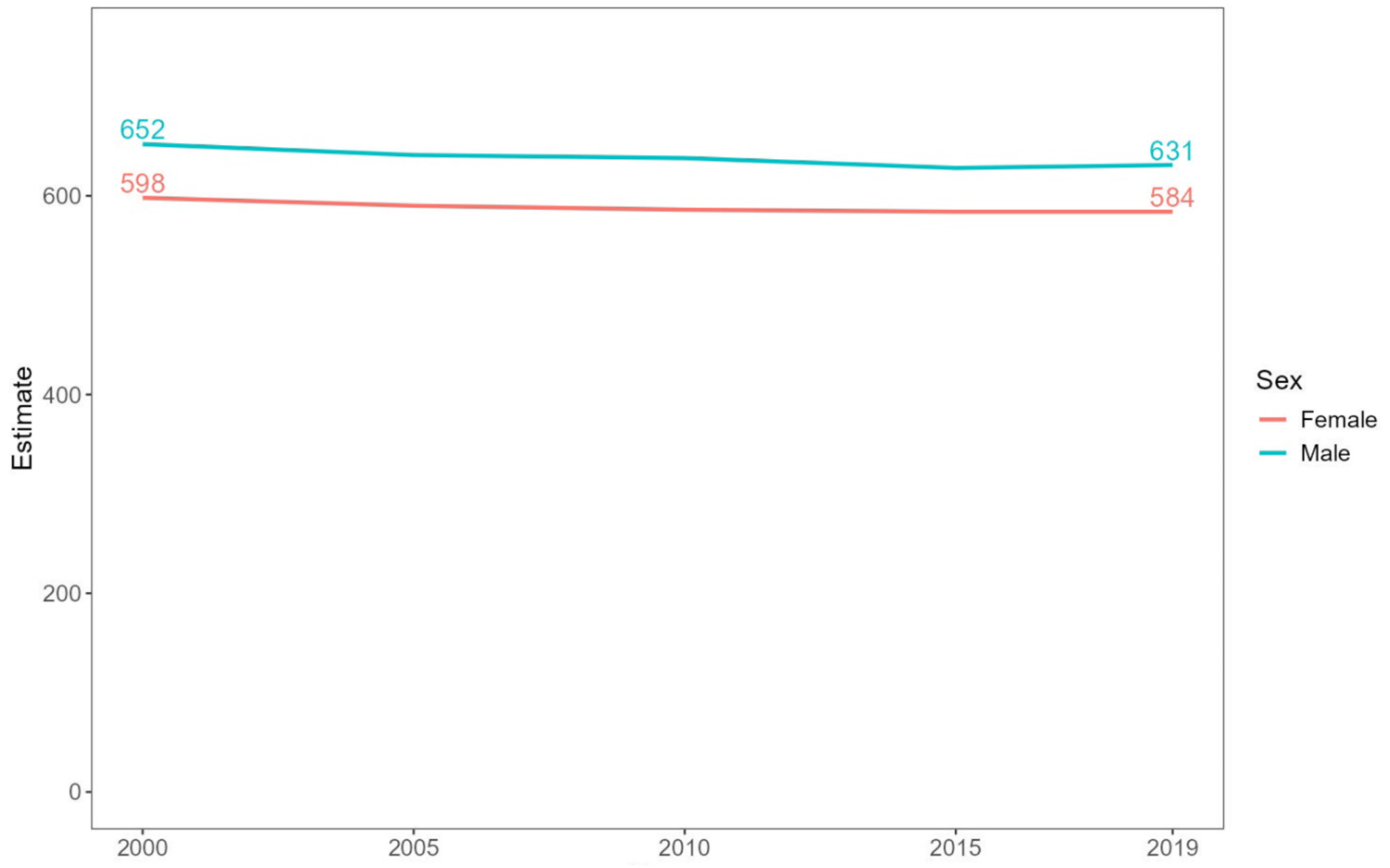
Cardiovascular Disease ASIR by sex (per 100 000 population) slowly decreasing and consistently higher in male.

Incidence of chronic respiratory disease was consistently higher in females compared UJ to males increasing from 665 cases per 100,000 population [95% CI 579-772] in 2000 to 729 cases per 100,000 population [95% CI 632-850] in 2019 (p-value 0.07) with a 0.41 annual percent increase while from 578 cases per 100,000 population [95% CI 507-671] in 2000 to 629 cases per 100,000 population [95% CI 543-736] in 2019 (p-value 0.17) in males with 0.37 annual percent increase (**figure 3**).

**Figure 3:**
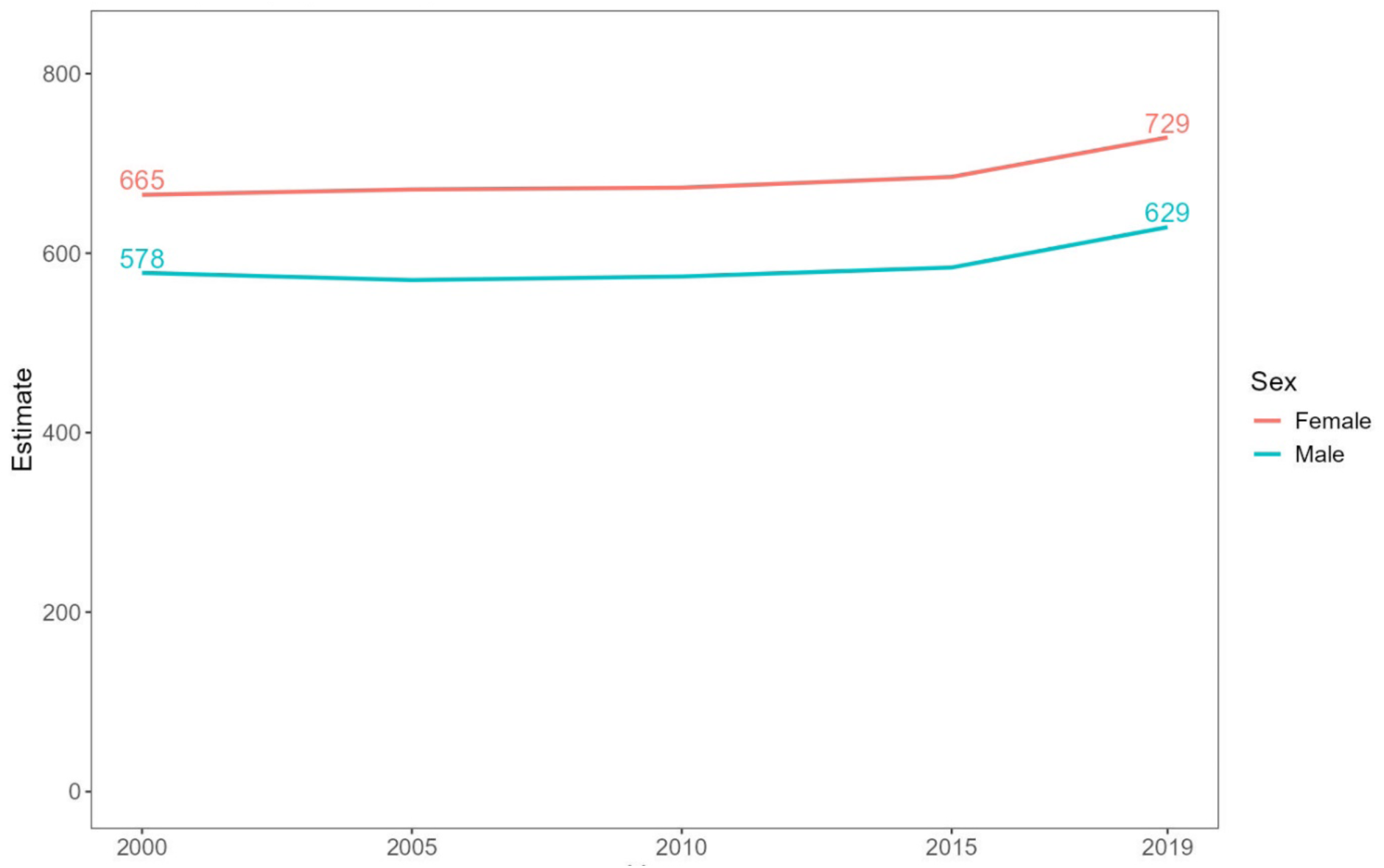
Chronic Respiratory Disease ASIR by sex per 100 000 population) increasing over time (p-value for trend 0.08) and consistently higher in females.

Throughout 2000-2019, the incidence rates of diabetes and kidney diseases in females were higher than those of males (**Figure 4**) and increased from 407 cases per 100,000 population [95% CI 382-434] in 2000 to 489 cases per 100,000 population [95% CI 455-525] in 2019 in females (p-value 0.04) with a 1.05 annual percent increase and from 397 cases per 100,000 population [95% CI 371-424] in 2000 to 464 cases per 100,000 population [95% CI 433-499] in 2019 in males (p-value 0.07) with a 0.85 annual percent increase. **Figure 5** shows comparisons of trends of different NCD in Senegal.

**Figure 4:**
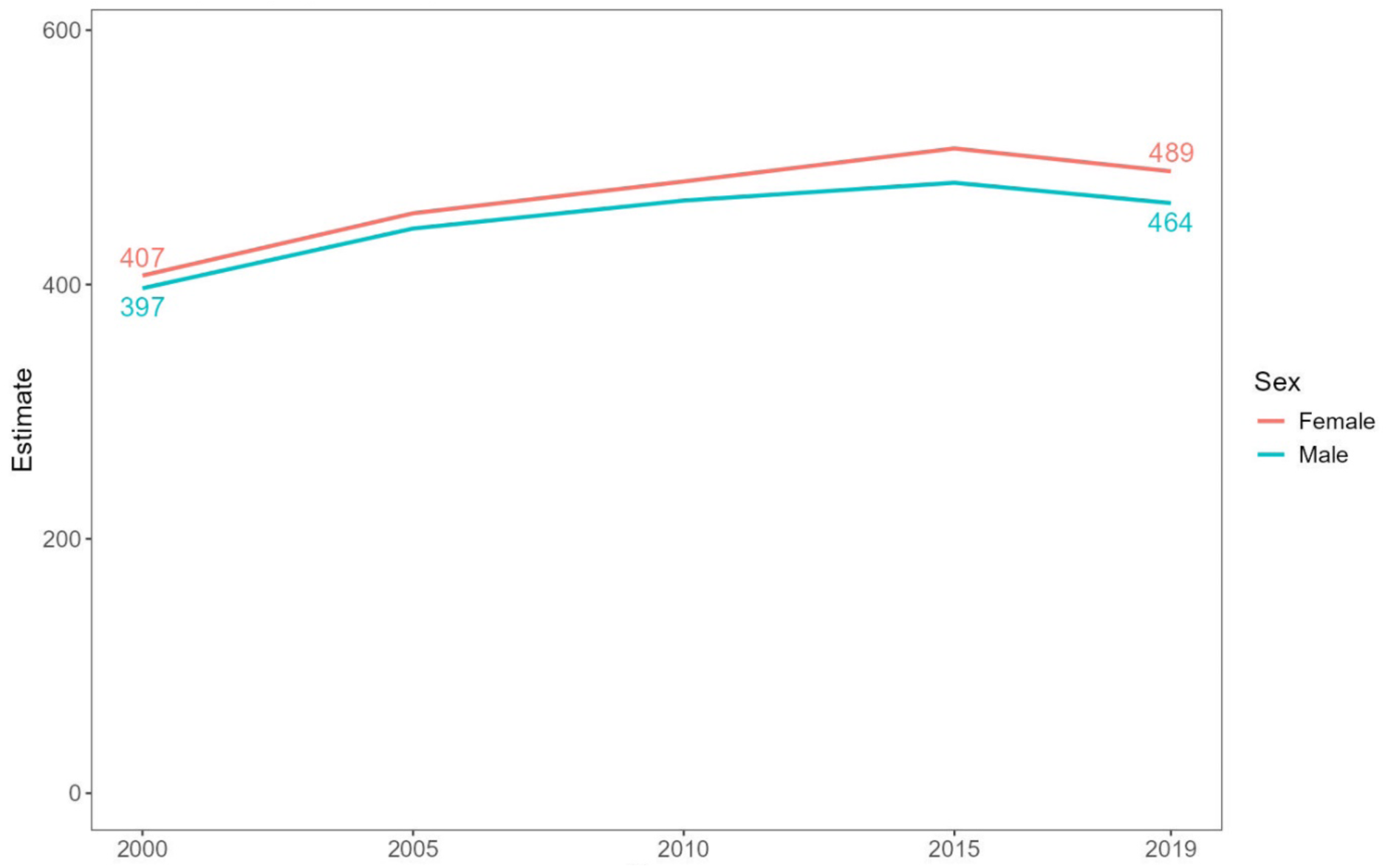
Diabetes and Chronic Kidney Disease ASIR by sex per 100 000 population) rising until 2015, followed by a decline, with rates remaining consistently higher in females.

**Figure 5:**
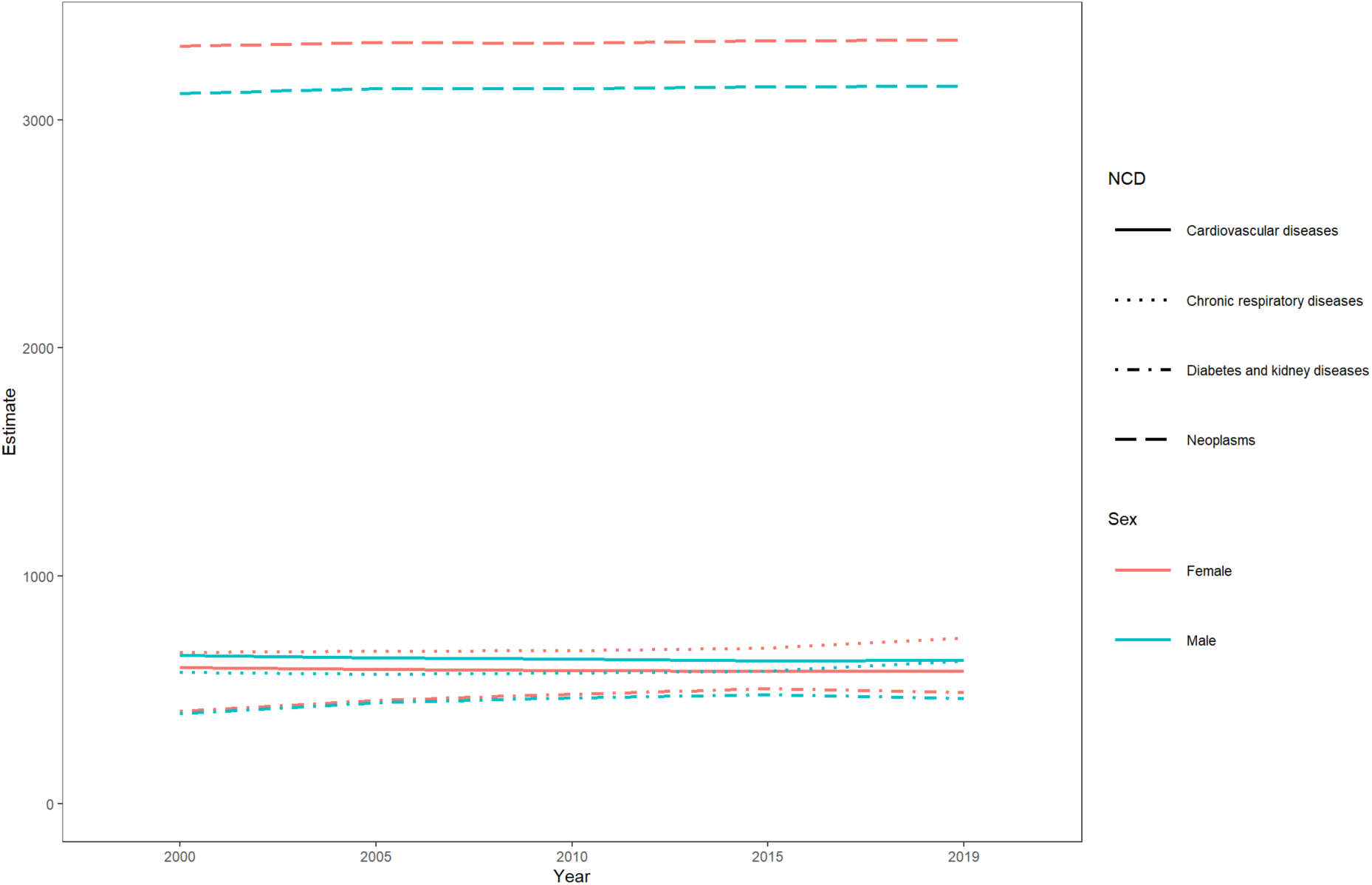
Comparison of non-communicable diseases subtypes ASIR by sex (per 100 000 population)

## DISCUSSION

### Key findings

We observed that NCD incidence in Senegal increased modestly between 2000 and 2019, with important variations by disease type and sex. Women experienced consistently higher overall NCD incidence than men throughout the study period, a pattern that held across most disease categories. The sex disparity was particularly pronounced for chronic respiratory diseases, where the rate of increase among women exceeded that of men. Cardiovascular disease presented a notable exception to this pattern, with men maintaining higher incidence rates despite declines in both sexes. While neoplasm incidence increased only marginally, diabetes and kidney disease showed the steepest rises among all NCD subtypes, suggesting an emerging epidemiological transition. These divergent trajectories across NCD subtypes and between sexes highlight the heterogeneous nature of the NCD epidemic in Senegal and underscore the need for targeted prevention strategies that account for both disease-specific and sex-specific risk factors.

The observed increase in NCD incidence is concerning and mirrors regional and global patterns consistent with the ongoing epidemiological transition. In West Africa, Ghana and Nigeria have reported similar increases in hypertension and diabetes, attributed to urbanisation, sedentary lifestyles, and dietary change.^21,22^ South Africa, with its high prevalence of obesity and smoking, has shown pronounced increases in NCD incidence rates. ^23^Senegal reached its epidemiological transition in 2014 (the point at which deaths from NCD outweighed deaths from communicable diseases).^24^ By 2019, deaths from NCD had risen to constitute almost half of all deaths.^24^

Multiple forces likely contribute to these trends in Senegal, including urbanisation, lifestyle change, and population ageing.^25,26^ The country is shifting from predominantly rural to increasingly urban settlements: in 1990, 77% of Senegalese lived in rural areas (vs 64.2% across lower-middle-income African countries), whereas by 2019 8.2 million people (52.3%) lived in rural areas (vs 50.4% average), with urban growth concentrated along the coastal zone, particularly Dakar.^24^ Rapid urbanisation has been associated with insecurity and environmental risks (pollution, flooding, erosion).^24^ Concurrently, cardiovascular risk factors, including hypertension, diabetes, smoking, physical inactivity, and high-sugar/high-fat diets, are rising.^5,7,27^ Health-system constraints may amplify these patterns: limited access to high-quality care, rural–urban disparities, treatment costs, and low insurance coverage.^7^

Incidence rates of cancers were consistently higher in females than males in our study. This pattern aligns with Global Cancer Observatory 2018 estimates for West Africa, which report higher age-standardised incidence in women than in men (122.0 vs 95.6 per 100,000) and a higher cumulative risk to age 75 (12.7% vs 10.1%).^28^ Differences in exposures and screening access, including Breast Cancer Awareness Month (“Pink Month”), may influence detection and incidence. Projections indicate steep increases in cancer incidence across all ages in SSA from 2010 to 2030.^29^ Additionally, cancers are one of the key NCD that contribute to high morbidity and mortality in SSA, including Senegal.^30^ Mortality rates are reported to be high, with specific challenges in Africa related to late-stage cancer presentation, lack of awareness, and limited access to screening and treatment.^29^

Despite the overall rise in NCDs, our CVD incidence estimates show a slight decline. Importantly, mortality trends do not necessarily track incidence, because they are influenced by case fatality, treatment access, and competing risks. Regional syntheses of CVD mortality and DALYs report large declines in many high- and some middle-income countries, whereas reductions in SSA have been smaller and uneven; in several analyses, absolute CVD deaths in SSA continued to grow as populations expanded and aged.^31,32^

The epidemiological transition in SSA has accelerated markedly, with the proportion of total disability-adjusted life years (DALYs) attributable to NCDs rising from approximately 18.6% in 1990 to 29.8% in 2017, driven substantially by cardiovascular disease.^33^ The absolute number of cardiovascular deaths in SSA increased by more than 50% during this period, with mortality occurring at younger ages than in other regions, thereby amplifying both disability burden and years of life lost.^34,35^ These patterns underscore critical gaps in cardiovascular health that require comprehensive approaches encompassing prevention, service strengthening, and equitable access to quality care. In Senegal, nascent interventions show promise: cardiac rehabilitation services, established with the country’s first outpatient centre in 2019, and patient education initiatives launched in 2018 have demonstrated short-term improvements in risk factor control and health literacy, though population-level impact remains to be established.^36,37^ The higher cardiovascular disease burden we observed among men aligns with regional patterns across SSA, where male predominance in cardiovascular incidence, DALYs, and mortality likely reflects both differential risk factor exposure, particularly tobacco use, and underlying biological susceptibilities.^38^

The rising incidence of chronic respiratory diseases in Senegal, particularly among women, reflects distinctive exposure patterns in West African settings. Women’s higher disease burden likely stems from disproportionate exposure to household air pollution from cooking with biomass fuels, a finding corroborated by local data demonstrating female predominance and associations between poor household ventilation and obstructive lung disease.^39^

These sex-specific patterns underscore the need for interventions targeting both household and ambient air pollution alongside tobacco control measures aligned with global evidence on respiratory disease prevention.^40^ The chronic respiratory disease burden in LMICs is further complicated by the legacy effects of tuberculosis, HIV-related lung disease, rapid urbanisation, and emerging occupational hazards.^41^ Across SSA, chronic respiratory diseases contribute substantially to mortality and disability-adjusted life years, with trajectories remaining concerning given persistent household air pollution exposure and incomplete implementation of tobacco control policies.^41^ The temporal pattern we observed for diabetes and kidney disease, with increases until 2015 followed by slight reductions, reflects the complex and evolving epidemiology of metabolic diseases in transitional settings.^42–45^

This trajectory parallels broader regional trends: countries with the highest malaria burden experienced rising age-standardised mortality from type 2 diabetes and chronic kidney disease between 1990 and 2019.^46^ In Senegal, urban areas show particularly high diabetes prevalence, while health system constraints limit both prevention efforts and long-term disease management.^7^ The ongoing nutrition transition associated with urbanisation, characterised by dietary shifts toward energy-dense, ultra-processed foods, continues to drive diabetes risk while simultaneously complicating dietary management for those with established disease.^8,47^

### Study limitations

This study relies on model-based national estimates from GBD 2019 accessed via the WHO HIDR rather than raw, country-level case counts. Precision therefore reflects the availability, quality, and representativeness of Senegal-specific input data; where inputs are sparse, GBD’s spatiotemporal modelling borrows strength from regional neighbours, which can attenuate country-specific signals. We analysed ASIR that were directly standardised to the GBD world standard population; comparisons with studies using alternative standards (such as WHO 2000-2025) require caution, as absolute levels may differ even when trends align. Because the data are aggregated at population level, we lacked key covariates (socioeconomic status, treatment access, subnational risk factors), precluding individual-level inference and limiting our capacity to explain within-country heterogeneity. Heterogeneity in the underlying source mix (surveys, civil registration, registries, facility data) and temporal variation in ascertainment or coding may introduce inconsistencies across years and settings, including possible under-ascertainment in rural or underserved areas.

Trend estimation is further constrained by the four quinquennial time points (2005, 2010, 2015, 2019) available in HIDR. With only four observations, joinpoint regression has low power to detect inflections (at most one joinpoint), so we emphasised the average annual percent change (AAPC) and provided a weighted log-linear sensitivity analysis. Uncertainty was handled by back-calculating standard errors from reported 95% uncertainty intervals; because GBD intervals arise from posterior draws and can be asymmetric, this approximation may misstate variability. Finally, there is no classical sample size for these estimates.

Future work should prioritise routinely collected, high-quality primary data (registries, strengthened civil registration and vital statistics, and subnational surveillance), incorporate draw-level analyses to propagate uncertainty more faithfully, and use annual GBD series where possible to enhance temporal resolution. Linking modelled outcomes to subnational and socioeconomic covariates would improve interpretability and policy relevance, particularly for targeting NCD prevention and care.

### Implications for public health

The present study highlights the critical need to have reliable epidemiological estimates of NCD to inform evidence-based prevention and control strategies. Accurate data allows policymakers to allocate resources effectively and empowers healthcare providers to deliver timely and appropriate interventions. Therefore, investing in robust surveillance systems and epidemiological research such as electronic health record and digital registries^48^ and integrated disease surveillance and response^49^, is crucial to develop a comprehensive understanding of the NCD landscape and guide the implementation of effective prevention and control measures tailored to the needs of our population. Addressing the increasing challenge of NCD requires the implementation of comprehensive strategies. These strategies should prioritize targeted interventions and emphasize the importance of modifying risk factors, early detection, and ensuring equal access to healthcare services while integrating approaches that reflect local dietary habits, social structures, and cultural norms. Health campaigns should promote healthier adaptations of traditional Senegalese cuisine, such as reducing excessive oil, salt, and sugar in popular dishes like “thieboudienne”, while encouraging nutrient-dense local foods like millet, fonio, and leafy greens. Additionally, community-based physical activity programs should incorporate culturally relevant activities, such as traditional dance (sabar, mbalax) or walking groups making exercise more engaging and accessible.

## CONCLUSION

Using harmonised, model-based estimates, we demonstrate that age-standardised incidence of NCDs in Senegal rose over the past two decades, with marked heterogeneity by sex and subtype. Neoplasms, chronic respiratory diseases, and diabetes with kidney disease increased, often disproportionately affecting women, while cardiovascular disease incidence declined modestly yet remained higher in men. These divergent trajectories and consistent sex differentials reveal that uniform NCD strategies fail to address the complex epidemiological reality in West African settings. Effective policy requires targeted, evidence-based interventions tailored to disease-specific and population-specific needs. For chronic respiratory diseases, this means accelerating adoption of clean cooking technologies and strengthening tobacco control. For diabetes and kidney disease, primary care systems must be equipped for early detection and long-term management within financially protected benefit packages. Cardiovascular prevention demands sex-responsive approaches that integrate risk factor control with accessible acute and post-acute care. Crucially, these interventions require robust surveillance infrastructure. Senegal and neighbouring countries need sustained investment in civil registration systems, disease-specific registries, and routine risk factor surveillance that generate open, analysable data with appropriate uncertainty quantification to track genuine epidemiological change.

The distinction between mortality and incidence trends is critical: declining death rates may mask rising disease occurrence if improved treatment extends survival without preventing new cases. Only through integrated monitoring of incidence, mortality, and care quality, coupled with responsive health systems that link prevention, primary care, and specialist services, can Senegal effectively address its growing NCD burden while advancing health equity. This comprehensive approach offers the most promising pathway to altering the NCD trajectory across sub-Saharan Africa.

## ABBREVIATION LIST

AAPC: Average annual percent change
APC: Annual percent change
ASIR: Age-standardised incidence rate(s)
CI / CIs: Confidence interval(s)
CODEm: Cause of Death Ensemble model
DALYs: Disability-adjusted life years
GBD: Global Burden of Disease (Study)
GHDx: Global Health Data Exchange
HIDR: Health Inequality Data Repository
HIV: Human immunodeficiency virus
IHME: Institute for Health Metrics and Evaluation
LMICs: Low- and middle-income countries
NCD(s): Non-communicable disease(s)
SSA: Sub-Saharan Africa
WHO: World Health Organization
DisMod-MR 2.1 (GBD): Bayesian disease model meta-regression platform

## Acknowledgments

The authors would like to acknowledge financial support for this work from the University of Pittsburgh Office of The Vice Provost for Faculty Diversity and Development and the Center for African Studies.

## Ethical approval

N/A no human subjects involved.

## Author Contributions

Conceptualization: N.D.G., M.B.J., M.M.K., J.T., T.L.G.-W., and B.G.

Formal analysis: J.T., D.K., and B.G.

Funding acquisition: T.L.G.-W.

Supervision: T.L.G.-W. and B.G.

Validation: T.L.G.-W. and B.G.

Visualization: J.T. and B.G.

Writing – original draft: N.D.G. and M.B.J.

Writing – review & editing: All authors (N.D.G., M.B.J., T.L.G.-W., M.M.K., M.A., I.M., A.T.A., J.T., D.K., G.S., J.L.C., E.L.P.S., M.J., and B.G.).

## Funding

No external funding

## Competing interests

There are no competing interests for any author.

## Patient consent for publication

Not required.

## Data availability statement

Data are publicly available from the World Health Organization’s Health Inequality Data Repository (HIDR)

## Notes

### Competing Interest Statement

The authors have declared no competing interest.

### Summary of Updates

This version of the manuscript has been revised to update formatting errors.

## REFERENCES

1. WHO. Global Status Report on Noncommunicable Diseases 2010.; 2011.

2. Shilton T, Champagne B, Blanchard C, Ibarra L, Kasesmup V. Towards a global framework for capacity building for non-communicable disease advocacy in low- and middle-income countries. Glob Health Promot. 2013;20(4_suppl):6–19. doi:10.1177/1757975913501208

3. Engelgau MM, Sampson UK, Rabadan-Diehl C, et al. Tackling NCD in LMIC Achievements and Lessons Learned from the NHLBI-UnitedHealth Global Health Centers of Excellence Program. Glob Heart. 2016;11(1):5–15. doi:10.1016/j.gheart.2015.12.016

4. Oni T, Unwin N. Why the communicable/non-communicable disease dichotomy is problematic for public health control strategies: implications of multimorbidity for health systems in an era of health transition. Int Health. 2015;7(6):390. doi:10.1093/INTHEALTH/IHV040

5. Mbaye A, Babaka K, Ngaïde AA, et al. Prevalence of cardiovascular risk factors in semi-rural area in Senegal. Ann Cardiol Angeiol (Paris). 2018;67(4):264–269. doi:10.1016/j.ancard.2018.04.005

6. Ndiaye AA, Tall AB, Gueye B, et al. A Cross-Sectional Survey on Non-Communicable Diseases and Risk Factors in the Senegalese Army. Health N Hav. 2016;08(14):1529–1541. doi:10.4236/health.2016.814151

7. Belue R, Elewonibi BM, Associate R. Non-Communicable Disease and Diabetes Screening in Community Settings in Low- and Middle-Income Countries: A Study in Senegal, West Africa. Vol 10.; 2017. https://scholarship.law.slu.edu/jhlp/vol10/iss2/5

8. Seck SM, Dia DG, Doupa D, et al. Diabetes Burden in Urban and Rural Senegalese Populations: A Cross-Sectional Study in 2012. Int J Endocrinol. 2015;2015. doi:10.1155/2015/163641

9. Lachat C, Otchere S, Roberfroid D, et al. Diet and Physical Activity for the Prevention of Noncommunicable Diseases in Low- and Middle-Income Countries: A Systematic Policy Review. PLoS Med. 2013;10(6). doi:10.1371/JOURNAL.PMED.1001465

10. Mayén AL, Marques-Vidal P, Paccaud F, Bovet P, Stringhini S. Socioeconomic determinants of dietary patterns in low- and middle-income countries: a systematic review. Am J Clin Nutr. 2014;100(6):1520–1531. doi:10.3945/AJCN.114.089029

11. World Health Organization 2024 data.who.int, Senegal [Country overview]. Accessed June 28, 2024. https://data.who.int/countries/686

12. Islam SMS, Purnat TD, Phuong NTA, Mwingira U, Schacht K, Fröschl G. Non Communicable Diseases (NCDs) in developing countries: A symposium report. Global Health. 2014;10(1):1–8. doi:10.1186/S12992-014-0081-9/METRICS

13. Fiseha T, Alemayehu E, Kassahun W, Adamu A, Gebreweld A. Factors associated with glycemic control among diabetic adult out-patients in Northeast Ethiopia. BMC Res Notes. 2018;11(1). doi:10.1186/S13104-018-3423-5

14. Lépine A, Le Nestour A. The determinants of health care utilisation in rural Senegal. J Afr Econ. 2013;22(1):163–186. doi:10.1093/JAE/EJS020

15. Islam SMS, Purnat TD, Phuong NTA, Mwingira U, Schacht K, Fröschl G. Non Communicable Diseases (NCDs) in developing countries: A symposium report. Global Health. 2014;10(1). doi:10.1186/S12992-014-0081-9

16. Kirkby K, Bergen N, Baptista A, Schlotheuber A, Hosseinpoor AR. Data Resource Profile: World Health Organization Health Inequality Data Repository. Int J Epidemiol. 2023;52(5):e253–e262. doi:10.1093/ije/dyad078

17. Vos T, Lim SS, Abbafati C, et al. Global burden of 369 diseases and injuries in 204 countries and territories, 1990–2019: a systematic analysis for the Global Burden of Disease Study 2019. The Lancet. 2020;396(10258):1204–1222. doi:10.1016/S0140-6736(20)30925-9

18. Joinpoint Regression Program, Version 5.3.0.0 - November 2024; Statistical Methodology and Applications Branch, Surveillance Research Program, National Cancer Institute. Accessed February 25, 2025. https://www.cancer.gov/research/resources/resource/90

19. Irimata K, Bastian B, Clarke T, Curtin S, Rui P. Guidance for Selecting Model Options in the National Cancer Institute Joinpoint Regression Software.; 2022. doi:10.15620/cdc:118050

20. Higgins JPT, Thomas J, Chandler J, et al. Cochrane Handbook for Systematic Reviews of Interventions. Wiley; 2019. doi:10.1002/9781119536604

21. Atun R, Jaffar S, Nishtar S, et al. Improving responsiveness of health systems to non-communicable diseases. Lancet. 2013;381(9867):690-697. doi:10.1016/S0140-6736(13)60063-X

22. Adeloye D, Basquill C, Aderemi A V., Thompson JY, Obi FA. An estimate of the prevalence of hypertension in Nigeria: a systematic review and meta-analysis. J Hypertens. 2015;33(2):230–242. doi:10.1097/HJH.0000000000000413

23. Samodien E, Abrahams Y, Muller C, Louw J, Chellan N. Non-communicable diseases - A catastrophe for South Africa. S Afr J Sci. 2021;117(5). doi:10.17159/SAJS.2021/8638

24. Enoch Randy Aikins (2024) Senegal. Senegal - ISS African Futures. Accessed May 18, 2024. https://futures.issafrica.org/geographic/countries/senegal/

25. WHO. WHO Noncommunicable diseases fact sheets. Internet. Accessed April 27, 2024. https://www.who.int/news-room/fact-sheets/detail/noncommunicable-diseases

26. Bigna JJ, Noubiap JJ. The rising burden of non-communicable diseases in sub-Saharan Africa. Lancet Glob Health. 2019;7(10):e1295–e1296. doi:10.1016/S2214-109X(19)30370-5

27. Seck SM, Guéye S, Tamba K, Ba I. Prevalence of chronic cardiovascular and metabolic diseases in senegalese workers: A cross-sectional study, 2010. Prev Chronic Dis. 2013;10(1). doi:10.5888/pcd10.110339

28. Bray F, Ferlay J, Soerjomataram I, Siegel RL, Torre LA, Jemal A. Global cancer statistics 2018: GLOBOCAN estimates of incidence and mortality worldwide for 36 cancers in 185 countries. CA Cancer J Clin. 2018;68(6):394–424. doi:10.3322/CAAC.21492

29. Dalal S, Beunza JJ, Volmink J, et al. Non-communicable diseases in sub-Saharan Africa: What we know now. Int J Epidemiol. 2011;40(4):885–901. doi:10.1093/ije/dyr050

30. Amuyunzu-Nyamongo M. Noncommunicable diseases, injuries, and mental health: the triple burden in Africa. Pan Afr Med J.NLM (Medline). 2022;43:167. doi:10.11604/pamj.2022.43.167.38392

31. World Heart Federation. World Heart Report 2023.; 2023. Accessed August 10, 2025. https://world-heart-federation.org/wp-content/uploads/World-Heart-Report-2023.pdf

32. Alhuneafat L, Ta’ani O Al, Tarawneh T, et al. Burden of cardiovascular disease in Sub-Saharan Africa, 1990–2019: An analysis of the Global Burden of Disease Study. Curr Probl Cardiol. 2024;49(6). doi:10.1016/j.cpcardiol.2024.102557

33. Gouda HN, Charlson F, Sorsdahl K, et al. Burden of non-communicable diseases in sub-Saharan Africa, 1990–2017: results from the Global Burden of Disease Study 2017. Lancet Glob Health. 2019;7(10):e1375–e1387. doi:10.1016/S2214-109X(19)30374-2

34. Yuyun MF, Sliwa K, Kengne AP, Mocumbi AO, Bukhman G. Cardiovascular Diseases in Sub-Saharan Africa Compared to High-Income Countries: An Epidemiological Perspective. Glob Heart. 2020;15(1):15. doi:10.5334/gh.403

35. Moran A, Forouzanfar M, Sampson U, Chugh S, Feigin V, Mensah G. The epidemiology of cardiovascular diseases in sub-saharan Africa: The global burden of diseases, injuries and risk factors 2010 study. Prog Cardiovasc Dis. 2013;56(3):234–239. doi:10.1016/j.pcad.2013.09.019

36. Ka MM, Mboup WN, Ndao SCT, et al. Low Cost Equipment and Short Duration Program Are Not Barriers to Good Outcomes of Cardiac Rehabilitation in Senegalese Patients with Coronary Artery Disease. World J Cardiovasc Dis. 2021;11(9):421–433. doi:10.4236/WJCD.2021.119039

37. Gaye ND, Ngaide AA, Aw F, et al. Patient Education in Cardiovascular Disease in Subsaharan Africa: The “Heart School” Pilot Project at Ipms/Cheikh Anta Diop University of Dakar, Senegal. Published online 2024. doi:10.2139/SSRN.4739601

38. Sun J, Qiao Y, Zhao M, Magnussen CG, Xi B. Global, regional, and national burden of cardiovascular diseases in youths and young adults aged 15–39 years in 204 countries/territories, 1990–2019: a systematic analysis of Global Burden of Disease Study 2019. BMC Med. 2023;21(1):1–15. doi:10.1186/S12916-023-02925-4/FIGURES/6

39. Thiam S, Daffe ML, Thiam K, et al. Impact Of Obstructive Lung Diseases (Asthma and COPD) due to indoor air pollution and poor room ventilation quality in Medina (Senegal). Journal of Toxicology and Health. 2023;10(1):1. doi:10.7243/2056-3779-10-1

40. Burney PGJ, Patel J, Newson R, Minelli C, Naghavi M. Global and Regional Trends in Chronic Obstructive Pulmonary Disease Mortality 1990-2010. Eur Respir J. 2015;45(5):1239. doi:10.1183/09031936.00142414

41. Soriano JB, Kendrick PJ, Paulson KR, et al. Prevalence and attributable health burden of chronic respiratory diseases, 1990–2017: a systematic analysis for the Global Burden of Disease Study 2017. Lancet Respir Med. 2020;8(6):585. doi:10.1016/S2213-2600(20)30105-3

42. Ogurtsova K, da Rocha Fernandes JD, Huang Y, et al. IDF Diabetes Atlas: Global estimates for the prevalence of diabetes for 2015 and 2040. Diabetes Res Clin Pract. 2017;128:40–50. doi:10.1016/J.DIABRES.2017.03.024

43. Mills KT, Xu Y, Zhang W, et al. A systematic analysis of worldwide population-based data on the global burden of chronic kidney disease in 2010. Kidney Int. 2015;88(5):950–957. doi:10.1038/KI.2015.230

44. Ong K, Stafford L, McLaughlin SB, et al. Global, regional, and national burden of diabetes from 1990 to 2021, with projections of prevalence to 2050: a systematic analysis for the Global Burden of Disease Study 2021. Lancet. Published online 2023. doi:10.1016/s0140-6736(23)01301-6

45. Bikbov B, Bikbov B, Purcell CA, et al. Global, regional, and national burden of chronic kidney disease, 1990–2017: a systematic analysis for the Global Burden of Disease Study 2017. The Lancet. Published online 2020. doi:10.1016/s0140-6736(20)30045-3

46. Li Z, Shi J, Li N, Wang M, Jin Y, Zheng Z jie. Temporal trends in the burden of non-communicable diseases in countries with the highest malaria burden, 1990– 2019: Evaluating the double burden of non-communicable and communicable diseases in epidemiological transition. Global Health. 2022;18(1). doi:10.1186/s12992-022-00882-w

47. Foley E, BeLue R. Identifying Barriers and Enablers in the Dietary Management of Type 2 Diabetes in M’Bour, Senegal. J Transcult Nurs. 2017;28(4):348–352. doi:10.1177/1043659616649028

48. Fraser HSF, Mugisha M, Remera E, et al. User Perceptions and Use of an Enhanced Electronic Health Record in Rwanda With and Without Clinical Alerts: Cross-sectional Survey. JMIR Med Inform. 2022;10(5):e32305. doi:10.2196/32305

49. Fall IS, Rajatonirina S, Yahaya AA, et al. Integrated Disease Surveillance and Response (IDSR) strategy: current status, challenges and perspectives for the future in Africa. BMJ Glob Health. 2019;4(4):e001427. doi:10.1136/BMJGH-2019-001427

